# Interventions designed to improve vaccination uptake: Scoping review of systematic reviews and meta-analyses - (version 1)

**DOI:** 10.1101/2022.01.26.22269890

**Authors:** CJ Heneghan, A Plüddemann, EA Spencer, J Brassey, EC Rosca, IJ Onakpoya, DH Evans, JM Conly, T Jefferson

## Abstract

**Background:** Resources to ensure high vaccination uptake differ widely across countries, but the best use of these precious resources is unclear. To better meet immunization programmes’ a pressing need to understand what works, particularly in low-resource settings, the World Health Organization commissioned a scoping view.

**Methods:** We conducted a scoping review of interventions designed to increase vaccination uptake, including systematic reviews and meta-analyses of interventional studies. We searched the following electronic databases: MEDLINE, Cochrane Database of Systematic Reviews, EMBASE, Epistemonikos, Google Scholar, LILACs and TRIP database until 01 July 2021 and hand-searched the reference lists of included articles. We included systematic reviews if they summarized studies with quantitative data on the impact on vaccine uptake for any age group. To assess review quality, we used a modified AMSTAR score. To evaluate the quality of the evidence in included reviews, we used the Grading of Recommendations Assessment, Development and Evaluation (GRADE).

**Results:** The final analysis set included 107 full-text reviews. Publication of reviews increased markedly over time, from seven reviews in 2010 to 38 reviews filtered in 2021. We conducted quality assessments for 72 reviews (132 outcomes). Based on the AMSTAR criteria, 40 included reviews (56%) received a quality rating of good, while the remaining 32 (44%) were of moderate quality. Only 13 reviews summarized data primarily for low- and middle-income countries (LMIC). The interventions were commonly multi-component, educational or reminder interventions; the description of intervention components was suboptimal and heterogeneous across most reviews. Effect estimates were available for 73 outcomes; in 52 (71%) of these, interventions led to statistically significant higher vaccine uptake compared with controls.

**Conclusions:** The literature has a large number of relevant systematic reviews on interventions to increase vaccine uptake, with an increased publication rate over time. However, problems with the definitions and the current reporting of vaccine uptake evidence make it difficult to determine what works best in low-resource settings.

## BACKGROUND

Vaccine uptake varies substantially by age, gender, ethnicity, geographical location, and socioeconomic status. Research has established that some of these differences are due to variations in vaccination’s behavioural and social drivers (BeSD). [1] In addition, the resources required to successfully promote the uptake of vaccines varies widely by country and income level. As a result, immunization programmes often struggle to achieve optimal coverage of the target population. [2]

Information on vaccine uptake can help vaccination programmes to understand what works for whom and in what settings. WHO seeks to complement its ongoing work on the measurement of BeSD with information on effective interventions to increase vaccine uptake. This information can help vaccination programmes to understand what works for whom and in what settings, particularly in low- and middle-income countries and other locations where resources are scarce.

Therefore, we undertook a scoping review of systematic reviews of published evidence on interventions designed to increase vaccine uptake across different populations and geographical regions to inform immunization programmes’ selection of interventions to promote vaccine uptake.

## Methods

We undertook a scoping review of interventions designed to increase vaccination uptake, based on methods developed in consultation with content experts at the World Health Organization. Our review focused on three topics:

- Interventions: any intervention designed to improve participation in vaccination;
- Outcomes: Uptake, Hesitancy, Disease risk appraisal, Confidence, Social norms, Provider recommendation, Availability; and
- Population: e.g. children, adolescents, adults and older adults (age 65+).

We conducted the review according to Preferred Reporting Items for Systematic Reviews and Meta-Analyses (PRISMA) guidelines. [3]

### Search strategy

We searched the following electronic databases: MEDLINE, Cochrane Database of Systematic Reviews, EMBASE, Epistemonikos, Google Scholar, LILACs and TRIP database (which covers guidelines and the grey literature) from 1 Jan 2010 until 01 July 2021 and hand-search the reference lists of included articles. The searches combined free and thesaurus search terms and keywords related to vaccine uptake and use sensitive search filters to focus on systematic reviews and meta-analyses. The search terms appear in Appendix 1. [4] We also search the bibliographies of retrieved systematic reviews and screen all titles and abstracts of retrieved citations for inclusion.

### Eligibility criteria

We included systematic reviews and meta-analyses of interventional studies that reported quantitative data on the impact on vaccine uptake for any age group. We included systematic reviews that primarily contained randomized controlled trials (RCTs); reviews were also included if they contained observational studies and quasi-experiments (including interrupted time series and before-and-after studies). However, we excluded reviews that assessed only vaccine efficacy or effectiveness and those that did not include any RCTs. We used the Jadad decision algorithm when there was more than one systematic review addressing the same or very similar therapeutic question in the same populations to select the most appropriate review. [5]; for instance, if multiple reviews addressed the same research question, we included the more recent, better quality and/or more comprehensive review. Two authors independently assessed review eligibility and reached a consensus over which review/meta-analysis to include.

### Data extraction

Three reviewers (CJH, EAS, IJO) through a consensus decided what data to extract from the included reviews. A reviewer extracted data from included reviews and a second reviewer independently checked the extraction. We reported outcomes by vaccine coverage, intervention type and population and created categories using an iterative process to extract outcomes. We extracted data on the population, study characteristics (e.g. the number of trials, location), the intervention as described by the authors, its comparator and the outcomes of interest and the year our search retrieved the publication (this could differ by electronic or print publication or in the bibliography it was retrieved from). Where two reviews covered the same intervention and outcome with overlapping studies, we selected the most relevant review (i.e. more comprehensive and up-to-date) for inclusion. One reviewer [EAS] extracted the data, and these were independently checked by a second reviewer [IJO]. Disagreements were resolved through consensus. Where a consensus could not be reached, a third reviewer [CJH] arbitrated.

### Quality assessment

To assess the quality of the included reviews we used the AMSTAR score and considered items 3 and 7 as essential for GRADE assessment: item 3, was a comprehensive literature search performed?; and item 7 was the scientific quality of the included studies assessed and documented? [6] We rated the quality of the evidence for the outcomes in the included reviews using the “Grade of Recommendations Assessment, Development and Evaluation” (GRADE). [7] We downgraded or upgraded the rating for the quality of the evidence, based on the amount of potential bias due to study design and other criteria specified in GRADE, and provide a summary of findings tables by the outcomes of interest. GRADE assessment was based on assessing the risk of bias and an evaluation of inconsistency, indirectness, and imprecision of the results and other factors See the Cochrane Handbook for further details on GRADE [8] and Appendix 4 for the GRADE assessments. Where the included reviews had already performed GRADE, we documented GRADE ratings as reported by the review authors. Two reviewers (EAS, IJO) performed (or documented) GRADE assessments for different sets of included reviews and then independently cross-checked each other’s assessments. We did not use other quality assessments by authors. Disagreements were resolved through discussion, and where necessary a third reviewer arbitrated (CJH).

### Outcomes of interest

We prioritized outcomes according to the WHO Handbook for Guideline Development [9] as high (critical for decision making in the context of the WHO BeSD Working Group), moderate (important for decision making) and low (not important for decision making). Vaccination uptake is high priority. Constructs in the WHO BeSD Framework are of moderate priority. Other constructs not in the framework are low priority (Table 1. Outcomes of interest and prioritization).

We present summary tables of the evidence by vaccine coverage, the specific population (i.e. n children, adolescents and older adults), and setting (e.g. healthcare, community) and also identify when a review separated findings primarily for LMICs.

We posted the protocol and findings (Appendix 1. Protocol_Vaccine Uptake_V1_2021.docx) in full on FigShare: https://figshare.com/s/5416371b9164af1ed716 and on MedXriv. See Interventions designed to improve vaccination uptake: Scoping review of systematic reviews and meta-analyses - protocol (version 1) medRxiv 2021.08.18.21262232; doi: https://doi.org/10.1101/2021.08.18.21262232)

## RESULTS

The literature searches identified 264 reviews for possible inclusion (see Figure 1. Vaccine Uptake Flow chart & Inclusion). Most reviews (240) came from the database search, and 24 came from reference searches. Reviews increased markedly over time: in 2010, only 7 reviews were published. However, by 2021, 38 reviews were published (see Figure 2). A further 129 reviews were excluded as they were not systematic or did not include RCTs or vaccine interventions, and 28 were further excluded as they were earlier versions or contained a lower number of RCTs (see Appendix 2).

We assessed 107 full-text reviews (Appendix 3). Of these, the most common vaccines studied were influenza (n=19) and HPV (n=12 reviews). We found four reviews for Hep B or C; Pertussis and Pneumococcal three reviews each; BCG two reviews and one each for MMR and Tetanus. The most common population included was children, with 14 reviews including childhood vaccines and eight including early childhood vaccines (See Table 2).

We defined 48 different interventions across the reviews reporting specific interventions (see Appendix 4). Reviews of multiple interventions (n=37) reported on 60 different interventions (see Appendix 5). These reviews most commonly included multi-component interventions, educational and/or reminder interventions. However, intervention reporting for these reviews was often suboptimal, meaning it was often difficult to discern and index the interventions.

We included 72 systematic reviews with 132 reported outcomes in the AMSTAR and GRADE assessment: 35 reviews were not assessed for the following reasons: no relevant outcomes assessed (n=14); no assessment of bias/study quality (n=11); overlapping/duplicate data of included more comprehensive/recent review (n=7); duplicate publication (same authors and studies, n=1) or superseded by an updated review (typically Cochrane, n=2).

### Quality Assessment (n=72 reviews)

Based on the AMSTAR criteria, 40 included reviews (56%) were rated to be of good quality, while the remaining 32 (44%) were of moderate quality (see Table 3 AMSTAR Score Vaccine Uptake Reviews). We conducted GRADE assessments for 72 reviews (132 outcomes). (See table 4. Vaccine Uptake Reviews). We excluded 35 reviews from the GRADE assessment for various reasons (see Figure 1 and Appendix 6. Excluded reviews from GRADE on full-text assessment).

### Outcomes

Effect estimates following meta-analysis of data were available for 73 outcomes. In 52 of these reviews, the interventions led to statistically significant higher vaccine uptake compared with controls.

Table 5 reports the Study Characteristics for these Vaccine Uptake SRs. We found 13 reviews primarily reporting LMIC populations assessed (Eze 2021, Yunusa 2021, Munk 2019, Lukusa 2018, Odendaal 2018, Bright 2017, Oyo-Ita 2016, Nelson 2016, Johri 2015, Jarrett 2015, Lassi 2015, Owusu-Addo 2014 and Bassani 2013); two reporting on Africa (Linde 2021 and Johnson 201, sub_Saharan Africa) and 37 reviews reporting exclusively on High-Income Countries (see table 5). The complete tracker is available online (see Appendix 7. Vaccine Uptake Tracker).

We applied GRADE to vaccine uptake in 114 of the 132 outcomes (86%) and nine knowledge outcomes (7%). The quality assessed through GRADE was moderate for 25 of 132 outcomes (19%), low for 79 outcomes (60%), and very low for 28 (21%). Generally, the main reasons for downgrading the quality of evidence were high heterogeneity and serious indirectness. Few outcomes that we applied GRADE to (13 of 73) showed considerable beneficial effects with the interventions, defined as a mean pooled effect estimate (risk ratio, odds ratio, or risk difference) of at least 2. However, 6 of 13 (46%) were rated as very low quality. Of the 18 reviews that showed moderate evidence for GRADE outcomes, seven were conducted exclusively in high-income settings, and six in LMICs.

## DISCUSSION

The literature searches identified a large number of reviews on interventions to increase vaccine uptake. Publication of reviews on vaccine uptake interventions have increased substantially over time. However, few reviews provided separate findings for LMICs, an important area for future research.

The quality of the reviews was good to moderate. The majority of intervention outcomes showed higher vaccine uptake compared with controls. The presence of some large effect estimates, however, should be treated with caution as half of these reviews were low quality. In addition, reviews generally included multi-component interventions, which previous research has shown to be reliably more effective than single-component interventions. Thus, many reviews in the current literature may not offer actionable insights about which specific components of multi-component interventions drive these intervention effects. Such information would help vaccination programmes more efficiently use scarce resources. We posted several appendices and tables on Figshare to make available the data for others to use (see List of Tables, Figures and Appendices).

Immunization remains an effective public health intervention. However, in 2020, 23 million children under the age of one missed out on basic vaccinations, the highest number since 2009. [10] The covid pandemic has seen substantial falls in immunisation rates, particularly in disadvantaged people and poorer countries.[11] Identifying effective strategies is, therefore, a pressing priority for policymakers globally.

Our review has several limitations. We identified what appeared to be at least five dozen different interventions based on the review authors’ descriptions, but the reporting was problematic. Because many reviews did not clearly define the interventions, an in-depth analysis would have required retrieving the primary studies, which was beyond the scope of this initial review. How the current reviews are produced and disseminated with no categorization or index of interventions, makes it difficult to assess what the actual interventions were and challenging to identify those intervention components that inform practice. The quality of the description of interventions in publications has been shown previously to be poor. The use of a template for intervention description and replication (the TIDieR checklists) is one potential way to improve the quality of reporting and aid implementation. [12]

### Implications for Policy and Research

We consider two areas where there are implications for improving practice and research: 1) improvements to the curation and dissemination of existing knowledge and 2) the generation of new knowledge.

### Existing knowledge

There is a need to improve the curation of knowledge for vaccine uptake interventions due to the chaotic overlapping nature of the existing reviews and an assessment of the gaps in knowledge. Nearly three-quarters of the reviews subject to GRADE reported beneficial impacts of interventions. Yet, there is still a need to better understand what works in what settings, and how programmes optimise the implementation of interventions dependent on their populations and the coverage to ensure optimal use of resources. This scoping review acts as a starting point for further analysis and consideration of how best to present the evidence for maximal impact. The next steps might involve the categorisation of interventions using the Measuring Behavioural and Social Drivers of Vaccination’ (BeSD) framework, the development of policy briefs that are digestible and readily searchable with links through to the original evidence

### New knowledge

The current definition and reporting of vaccine uptake interventions have an apparent problem. Many reviews use vague or no description of the interventions, and therefore, a taxonomy of interventions and standard definitions is required. Reviews and primary studies are also required in focused areas to fill gaps in knowledge particularly in LMICs for priority vaccines.

## CONCLUSION

We found a large number of relevant systematic reviews of vaccine uptake interventions, with an increasing rate of publication over time. However, problems with the definitions and the current reporting of vaccine uptake evidence can make it difficult to determine what works in what settings, dependent on resources. This literature would greatly benefit from better curation of knowledge to target resources and efficiently improve vaccine uptake.

## Data Availability

All data included in the review are provided in the Appendices, tables and text and made available at Figshare: https://figshare.com/s/5416371b9164af1ed716 files available to download)

## List of Tables, Figures and Appendices

for Interventions designed to improve vaccination uptake

Scoping review of systematic reviews and meta-analyses

(Reference to Figshare: https://figshare.com/s/5416371b9164af1ed716 10.6084/m9.figshare.15200286 files available to download)

| Appendices |  |
| --- | --- |
| Appendix 1. | Protocol_Vaccine Uptake_V1_2021.docx |
| Appendix 2. | Excluded Discordant Review references |
| Appendix 3. | Included References for GRADE Assessment (n=107) |
| Appendix 4. | Glossary of Intervention definitions |
| Appendix 5. | Multiple Intervention components across 37 SRs |
| Appendix 6. | Excluded reviews from GRADE (on full-text assessment) |
| Appendix 7. | Vaccine Uptake Tracker |
| Figures |  |
| Figure 1. | Vaccine Uptake Flow chart & Inclusion |
| Figure 2. | Vaccine Uptake Flow chart by year |
| Tables |  |
| Table 1. | Outcomes of Interest and Prioritization |
| Table 2. | Summary of Intervention coverage and populations |
| Table 3. | AMSTAR Score Vaccine Uptake Reviews |
| Table 4. | GRADE Tables Vaccine Uptake Reviews |
| Table 5. | Study Characteristics Vaccine Uptake SRs |

## Authors’ contributions

All authors contributed in equal part to the conceptualization and development of the content. TJ and CH wrote the first draft and edited this version. All authors contributed to the subsequent drafts and approved the final version. The authors acknowledge the substantial intellectual contribution of Dr Lisa Menning, Dr Julie Leask, and the WHO BeSD working group in conceptualizing the research question, outcomes, and intervention types.

## Conflict of interest statements

TJ received a Cochrane Methods Innovations Fund grant to develop guidance on using regulatory data in Cochrane reviews (2015 to 2018). From 2014 to 2016, he was a member of three advisory boards for Boehringer Ingelheim. TJ was a member of an independent data monitoring committee for a Sanofi Pasteur clinical trial on an influenza vaccine. Market research companies occasionally interview TJ about phase I or II pharmaceutical products for which he receives fees (current). TJ was a member of three advisory boards for Boehringer Ingelheim (2014 to 16). TJ was a member of an independent data monitoring committee for a Sanofi Pasteur clinical trial on an influenza vaccine (2015 to 2017). TJ is a relator in a False Claims Act lawsuit on behalf of the United States that involves sales of Tamiflu for pandemic stockpiling. If resolved in the United States favour, he would be entitled to a percentage of the recovery. TJ is coholder of a Laura and John Arnold Foundation grant for the development of a RIAT support centre (2017 to 2020) and Jean Monnet Network Grant, 2017 to 2020 for The Jean Monnet Health Law and Policy Network. TJ is an unpaid collaborator to the Beyond Transparency in Pharmaceutical Research and Regulation led by Dalhousie University and funded by the Canadian Institutes of Health Research (2018 to 2022). TJ consulted for Illumina LLC on next-generation gene sequencing (2019 to 2020). TJ was the consultant scientific coordinator for the HTA Medical Technology programme of the Agenzia per I Servizi Sanitari Nazionali (AGENAS) of the Italian MoH (2007 to 2019). TJ is Director Medical Affairs for BC Solutions, a market access company for medical devices in Europe. TJ was funded by NIHR UK and the World Health Organization (WHO) to update Cochrane review A122, Physical Interventions to interrupt the spread of respiratory viruses. Oxford University funds TJ to carry out a living review on the transmission epidemiology of COVID 19. Since 2020, TJ receives fees for articles published by The Spectator and other media outlets. TJ is part of a review group carrying out a Living rapid literature review on the modes of transmission of SARS CoV 2 (WHO Registration 2020/1077093 0). He is a member of the WHO COVID 19 Infection Prevention and Control Research Working Group, for which he receives no funds. TJ is funded to co-author rapid reviews on the impact of Covid restrictions by the Collateral Global Organisation.

CJH holds grant funding from the NIHR, the NIHR School of Primary Care Research, the NIHR BRC Oxford and the World Health Organization for a series of Living rapid reviews on the modes of transmission of SARs CoV 2, reference WHO registration No2020/1077093, and to carry out a scoping review of systematic reviews of interventions to improve vaccination uptake, reference WHO Registration 2021/1138353-0. He has received financial remuneration from an asbestos case and given legal advice on mesh and hormone pregnancy tests cases. He has received expenses and fees for his media work, including occasional payments from BBC Radio 4 Inside Health and The Spectator. He receives expenses for teaching EBM and is also paid for his GP work in NHS out of hours (contract Oxford Health NHS Foundation Trust). He has also received income from the publication of a series of toolkit books and appraising treatment recommendations in non-NHS settings. He is the Director of CEBM, an NIHR Senior Investigator and an advisor to Collateral Global.

DHE holds grant funding from the Canadian Institutes for Health Research and Li Ka Shing Institute of Virology relating to the development of Covid 19 vaccines and the Canadian Natural Science and Engineering Research Council concerning Covid 19 aerosol transmission. He is a recipient of World Health Organization and Province of Alberta funding which supports the provision of BSL3 based SARS CoV 2 culture services to regional investigators. He also holds public and private sector contract funding relating to the development of poxvirus based Covid 19 vaccines, SARS CoV 2 inactivation technologies, and serum neutralization testing.

JMC holds grants from the Canadian Institutes for Health Research on acute and primary care preparedness for COVID 19 in Alberta, Canada and was the primary local Investigator for a Staphylococcus aureus vaccine study funded by Pfizer, for which all funding was provided only to the University of Calgary. He is a co-investigator on a WHO funded study using integrated human factors and ethnography approaches to identify and scale innovative IPC guidance implementation supports in primary care with a focus on low resource settings and using drone aerial systems to deliver medical supplies and PPE to remote First Nations communities during the COVID 19 pandemic. He also received support from the Centers for Disease Control and Prevention (CDC) to attend an Infection Control Think Tank Meeting. He is a member of the WHO Infection Prevention and Control Research and Development Expert Group for COVID 19 and the WHO Health Emergencies Programme (WHE) Ad hoc COVID 19 IPC Guidance Development Group, both of which provide multidisciplinary advice to the WHO, for which no funding is received and from which no funding recommendations are made for any WHO contracts or grants. He is also a member of the Cochrane Acute Respiratory Infections Group.

JB is a major shareholder in the Trip Database search engine (www.tripdatabase.com) as well as being an employee. In relation to this work, Trip has worked with a large number of organizations over the years; none have any links with this work. The main current projects are with AXA and Collateral Global.

ECR was a member of the European Federation of Neurological Societies(EFNS) / European Academy of Neurology (EAN) Scientist Panel, Subcommittee of Infectious Diseases (2013 to 2017). Since 2021, she is a member of the International Parkinson and Movement Disorder Society (MDS) Multiple System Atrophy Study Group, the Mild Cognitive Impairment in Parkinson Disease Study Group, and the Infection Related Movement Disorders Study Group. She was an External Expert and sometimes Rapporteur for COST proposals (2013, 2016, 2017, 2018, 2019) for Neurology projects. She is a Scientific Officer for the Romanian National Council for Scientific Research.

IJO, EAS, and AP have no interests to disclose.

## Ethics committee approval

No ethics approval was necessary.

